# Routine measurement of serum procalcitonin allows antibiotics to be safely withheld in patients admitted to hospital with SARS-CoV-2 infection

**DOI:** 10.1101/2020.06.29.20136572

**Authors:** Emma J. Williams, Luke Mair, Thushan I. de Silva, Dan J. Green, Philip House, Kay Cawthron, Christopher Gillies, James Wigfull, Helena Parsons, David G. Partridge

## Abstract

**Background:** It can be a diagnostic challenge to identify COVID-19 patients without bacterial co-infection in whom antibiotics can be safely stopped. We sought to evaluate the validity of a guideline that recommends withholding antibiotics in patients with a low serum procalcitonin (PCT).

**Methods:** We retrospectively collected 28-day outcome data on patients admitted to Sheffield Teaching Hospitals NHS Foundation Trust, UK, between 5 March and 15 April 2020, with a positive SARS-CoV-2 polymerase chain reaction (PCR) and PCT within 48 hours of diagnosis. PCT was considered negative if ≤0.25ng/ml and positive if >0.25ng/ml. Primary outcomes included antibiotic consumption, mortality, intensive care admission and length of hospital stay.

**Results:** 368 patients met the inclusion criteria; 218 (59%) had a negative PCT and 150 (41%) positive. At 48 hours post-diagnosis, 73 (33%) of those with a negative PCT were receiving antimicrobials compared to 126 (84%) with a positive PCT (p<0.001), with a corresponding reduction in antimicrobial usage over 28 days (median DDD of 3.0 vs 6.8 (p<0.001); median DOT 2 vs 5 days (p<0.001) between the negative and positive PCT groups.) In the negative PCT group, there were fewer deaths (62 (28%) vs. 54 (36%), (p=0.021)) and critical care admissions (19 (9%) vs. 28 (19%), (p=0.007)) than in the positive PCT group. Median length of hospital stay was 8.7 and 9 days in the negative and positive PCT groups respectively.

**Conclusions:** Procalcitonin is a valuable tool in the assessment of patients with SARS-CoV-2 infection, safely reducing the potential burden of unnecessary antibiotic usage.

## BACKGROUND

In patients with COVID-19, the presentation of fever, tachypnoea and hypoxia, together with lung infiltrates on chest imaging and a frequent rise in biomarkers such as C-reactive protein (CRP) [1] presents a challenge to rational use of antimicrobials, as it is difficult to confidently identify or exclude bacterial co-infection. Rates of true bacterial co-infection are estimated to be only 7 to 14% [2-4]. Despite this, the International Severe Acute Respiratory and emerging Infections Consortium (ISARIC) World Health Organization (WHO) Clinical Characterisation Protocol UK (CCP-UK) study has shown that over 80% of patients with COVID-19 receive antibiotic treatment [5]. The Surviving Sepsis Campaign recommends empiric antibacterial therapy in its guidelines for the management of COVID-19 in critically ill adults [6] and the National Institutes of Health treatment guidelines suggest that, in the absence of evidence to the contrary, some expert clinicians advise administration of broad spectrum antimicrobials to all patients with moderate or severe hypoxia [7]. Unnecessary antibiotic usage could result in prolonged inpatient stay and adverse effects and optimising antibiotic use is part of national and global efforts to reduce the spread of antimicrobial resistance [8, 9]. Strategies to accurately identify patients with COVID-19 who do not have bacterial co-infection are urgently required [10]. Recent National Institute for Health and Care Excellence (NICE) guidance on pneumonia in the context of COVID-19 has recommended further research into the use of procalcitonin (PCT) for this purpose [4]. Furthermore, the co-chairs of the American Thoracic Society and Infectious Diseases Society of America Guideline for Treatment of Adults with Community Acquired Pneumonia (ATS) have endorsed the use of the assay to guide antibiotic usage [11].

Unlike other biomarkers such as CRP, PCT is not thought to be routinely raised in COVID-19 [12-15], although data on its utility in this context are scarce. A recent meta-analysis showed that COVID-19 patients with increased PCT values had a nearly 5-fold higher risk of severe infection, thought to be due to the presence of bacterial co-infection [16]. PCT has been shown to have a greater impact on antibiotic exposure than standard stewardship strategies alone [17], however this hypothesis remains untested in the setting of COVID-19. We set out to evaluate whether PCT use had an impact on i) antibiotic usage and ii) outcomes in patients with confirmed COVID-19 at a large NHS Foundation Trust Hospital in the United Kingdom (UK).

## METHODS

### Study Design, Study Site and Population

This retrospective observational study was undertaken at two sites of Sheffield Teaching Hospitals NHS Foundation Trust (STHNHFT; Royal Hallamshire Hospital and Northern General Hospital) with a combined total of 1700 beds. Patients diagnosed between 5 March and 15 April 2020 were included in the study, allowing assessment of 28-day outcome in all patients by the time of analysis. Those diagnosed prior to 5th March were excluded as at that stage COVID-19 was managed as a high consequence infectious disease and patients were admitted regardless of symptom severity, making the group unrepresentative of those admitted later in the epidemic. The enrolment end date of 15th April was before mandatory SARS-CoV-2 screening of all patients admitted to hospital was introduced.

Eligible patients were aged ≥ 18 years old, admitted to STHNFT, with both a positive SARS-CoV-2 reverse-transcriptase polymerase chain reaction (RT-PCR) result on nose and/or throat swabs and/or deep respiratory samples, and a PCT assay undertaken within 48 hours of collection of the SARS-CoV-2 sample. RT-PCR was performed using an in-house assay targeting E and RdRP genes [18]. Patients with both community and nosocomial acquisition of COVID-19 were included. STHNFT guidelines recommended that antibiotics could be withheld in COVID-19 patients with a PCT value of ≤0.25ng/ml unless felt necessary by a senior clinician, as concomitant bacterial infection is unlikely in such patients [19].

The study was granted approval by the STHNFT Clinical Effectiveness Unit (Ref: 9863)

### Data Collection and Outcomes

Demographic and clinical characteristics of patients were drawn from existing laboratory, pharmacy and clinical databases and from examination of physical and electronic patient notes. Data were entered into an electronic case report form (Access 2010, Microsoft, Redmond, WA, USA) including age, sex, ethnicity, height, weight, comorbidities, and antibiotic allergies.

28-day outcome was recorded as discharged, still in hospital or died. Data on readmissions was also collected and the duration of readmissions up to day 28 was incorporated into length of stay data for each patient. Adverse events within the 28 days of follow up were recorded including: Hospital-acquired pneumonia/ventilator-associated pneumonia (HAP/VAP), *Clostridioides difficile* infection, Meticillin-resistant *Staphylococcus aureus* (MRSA) acquisition and isolation of an extended-spectrum beta-lactamase (ESBL) or AmpC beta-lactamase producing organism from a clinical sample. HAP/VAP was defined as commencement of a new antibacterial agent for a presumed chest source at least 48 hours after COVID-19 diagnosis, in conjunction with either new elevation of white blood cell count or neutrophils or positive sputum culture for an organism other than low virulence oral flora. This definition was used as other components of the Infectious Diseases Society of America (IDSA)/ American Thoracic Society (ATS) definition of HAP/VAP (new lung infiltrates, fever and decline in oxygenation) would be difficult to discern in the context of COVID-19 [20].

Antibiotic usage was recorded for the 28-day follow up period including antibiotic agent, duration, and WHO defined daily doses (DDD). Where different DDDs are defined for oral and parenteral preparations, the parenteral figure was used for calculations to avoid inappropriate weighting by route of administration for some commonly used agents (e.g. clarithromycin) which would not be relevant from a stewardship perspective [21]. Prescribing rates were calculated using DDD/evaluable day for which data of patients who died during the 28-day follow up period were censored from the date of death. Days of treatment (DOT) were defined as the number of days in the 28-day period for which antibiotics were prescribed.

### Statistical Analysis

All values from patients meeting eligibility criteria were summarised using the most appropriate form, either using frequency/percentages, or medians with IQR (Inter-Quartile Range). In the scenario where there are multiple zeros values, and the IQR becomes uninformative with values of 0.00 to 0.00, the 10^th^ and 90^th^ centile are reported instead. Differences between demographics were analysed with the suitable significance test, depending on whether parametric assumptions were met as is detailed in the tables. To investigate the relationship between PCT positivity and total DDD, antibiotic receipt at 48 hours post-diagnosis and meropenem prescription, linear and logistic regression models were explored adjusting for demographic confounders (age, sex, ethnicity and comorbidities). Regression model assumptions were examined, and appropriate adjustments made. All p-values presented were two-sided, and estimates provided with a 95% confidence interval (CI). All statistical analyses were performed in Stata version 16.1 (StataCorp 2019. Stata Statistical Software: Release 16. College Station, TX: StataCorp LLC.)

## RESULTS

### Study Population

A total of 368 patients met the eligibility criteria and were included in the analysis; overall 60% were male, with a median age of 75. Of these, 218 (59%) had a PCT level of ≤0.25 (negative) and 150 (41%) had a level of >0.25 (positive). Patient demographics and comorbidities stratified by PCT results are available in Table 1. There was no significant difference in demographics between the two groups in terms of age, sex, BMI or ethnicity. Comorbidities between the two groups were also similarly distributed with the exception of malignancy, which was more common in the PCT negative group. There were no pregnant women in the cohort.

**Table 1.**
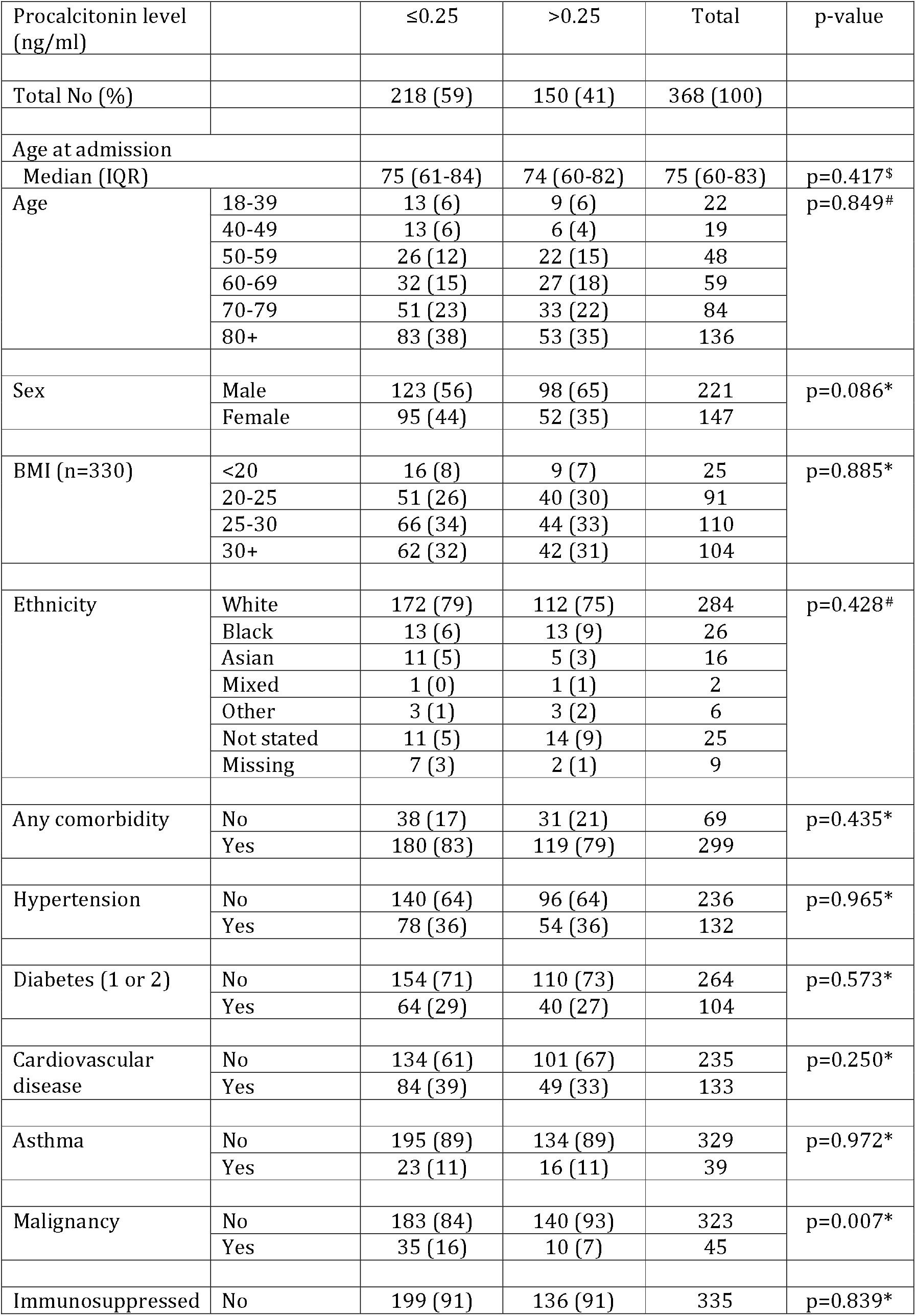
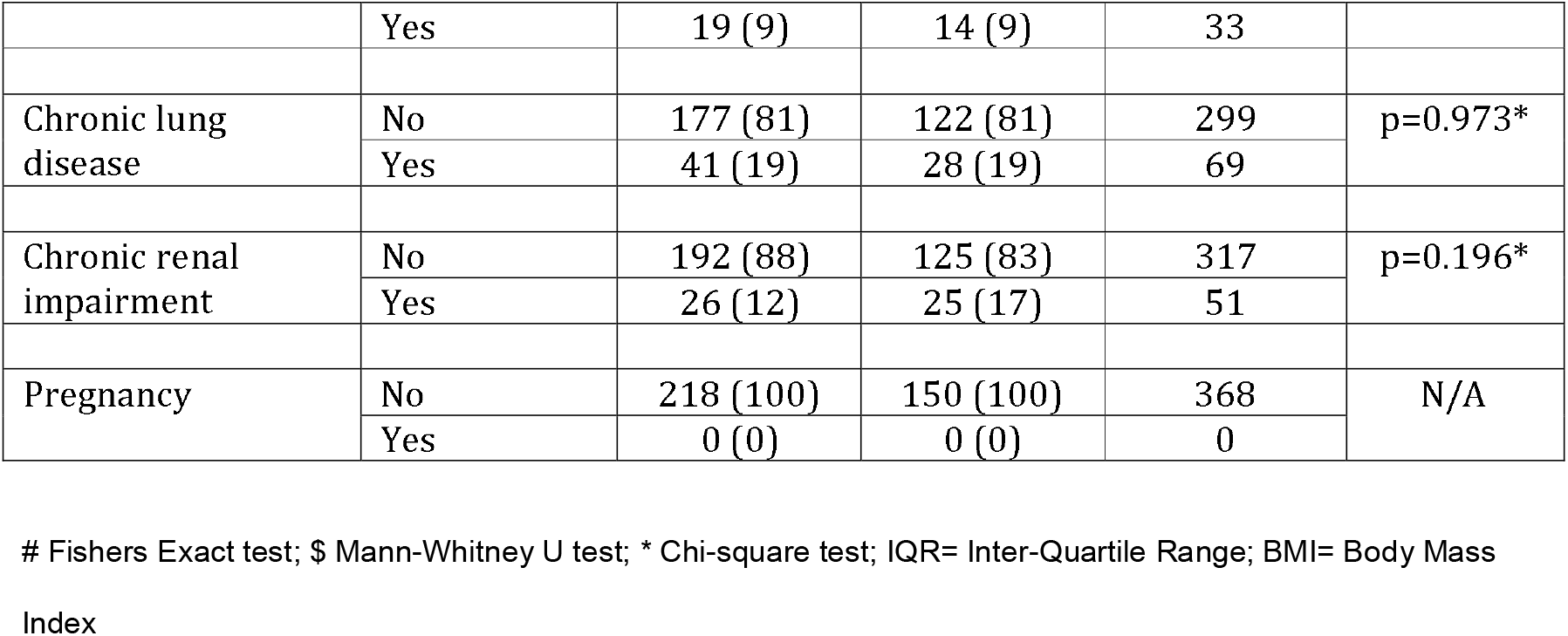
Baseline demographics details of patients stratified by procalcitonin level.

| Procalcitonin level (ng/ml) |  | ≤0.25 | >0.25 | Total | p-value |
| --- | --- | --- | --- | --- | --- |
| Total No (%) |  | 218 (59) | 150 (41) | 368 (100) |  |
| Age at admission |  |  |  |  |  |
| Median (IQR) | | 75 (61-84) | 74 (60-82) | 75 (60-83) | p=0.417\$ |
| Age | 18-39 | 13 (6) | 9 (6) | 22 | p=0.849# |
|  | 40-49 | 13 (6) | 6 (4) | 19 |  |
|  | 50-59 | 26 (12) | 22 (15) | 48 |  |
|  | 60-69 | 32 (15) | 27 (18) | 59 |  |
|  | 70-79 | 51 (23) | 33 (22) | 84 |  |
|  | 80+ | 83 (38) | 53 (35) | 136 |  |
| Sex | Male | 123 (56) | 98 (65) | 221 | p=0.086* |
|  | Female | 95 (44) | 52 (35) | 147 |  |
| BMI (n=330) | <20 | 16 (8) | 9 (7) | 25 | p=0.885* |
|  | 20-25 | 51 (26) | 40 (30) | 91 |  |
|  | 25-30 | 66 (34) | 44 (33) | 110 |  |
|  | 30+ | 62 (32) | 42 (31) | 104 |  |
| Ethnicity | White | 172 (79) | 112 (75) | 284 | p=0.428# |
|  | Black | 13 (6) | 13 (9) | 26 |  |
|  | Asian | 11 (5) | 5 (3) | 16 |  |
|  | Mixed | 1 (0) | 1 (1) | 2 |  |
|  | Other | 3 (1) | 3 (2) | 6 |  |
|  | Not stated | 11 (5) | 14 (9) | 25 |  |
|  | Missing | 7 (3) | 2 (1) | 9 |  |
| Any comorbidity | No | 38 (17) | 31 (21) | 69 | p=0.435* |
|  | Yes | 180 (83) | 119 (79) | 299 |  |
| Hypertension | No | 140 (64) | 96 (64) | 236 | p=0.965* |
|  | Yes | 78 (36) | 54 (36) | 132 |  |
| Diabetes (1 or 2) | No | 154 (71) | 110 (73) | 264 | p=0.573* |
|  | Yes | 64 (29) | 40 (27) | 104 |  |
| Cardiovascular disease | No | 134 (61) | 101 (67) | 235 | p=0.250* |
|  | Yes | 84 (39) | 49 (33) | 133 |  |
| Asthma | No | 195 (89) | 134 (89) | 329 | p=0.972* |
|  | Yes | 23 (11) | 16 (11) | 39 |  |
| Malignancy | No | 183 (84) | 140 (93) | 323 | p=0.007* |
|  | Yes | 35 (16) | 10 (7) | 45 |  |
| Immunosuppressed | No | 199 (91) | 136 (91) | 335 | p=0.839* |
|  | Yes | 19 (9) | 14 (9) | 33 |  |
| Chronic lung disease | No | 177 (81) | 122 (81) | 299 | p=0.973* |
|  | Yes | 41 (19) | 28 (19) | 69 |  |
| Chronic renal impairment | No | 192 (88) | 125 (83) | 317 | p=0.196* |
|  | Yes | 26 (12) | 25 (17) | 51 |  |
| Pregnancy | No | 218 (100) | 150 (100) | 368 | N/A |
|  | Yes | 0 (0) | 0 (0) | 0 |  |

### Compliance with Guideline

Of those patients with a negative PCT, 73 (33%) were on antibiotics 48 hours after their COVID-19 diagnosis compared to 126 (84%) with a positive PCT (p<0.001) suggesting good compliance with the guideline (Table 2).

**Table 2.** Antibiotic use and 28 day outcomes in patients stratified by procalcitonin level.

| Procalcitonin level (ng/ml) |  | ≤0.25 | >0.25 | Total | p-value |
| --- | --- | --- | --- | --- | --- |
| Total (%) |  | 218 (59) | 150 (41) | 368 (100) |  |
| **Clinical Outcomes** |  |  |  |  |  |
| 28 day outcome | Died | 62 (28) | 54 (36) | 116 (32) | p=0.021<sup>#</sup> |
|  | Discharged | 147 (67) | 82 (55) | 229 (62) |  |
|  | Still in hospital | 9 (4) | 14 (9) | 23 (6) |  |
| Intubated | No | 207 (95) | 129 (86) | 336 (91) | p=0.004<sup>#</sup> |
|  | Yes | 11 (5) | 21 (14) | 32 (9) |  |
| Admitted to ITU | No | 199 (91) | 122 (81) | 321 (87) | p=0.007<sup>#</sup> |
|  | Yes | 19 (9) | 28 (19) | 47 (13) |  |
| Length of stay |  |  |  |  |  |
| Median (IQR) | | 8.7 (4.9-15.3) | 9.0 (5.9-18.8) | 8.9 (5.3-16.1) | p=0.054<sup>\$</sup> |
| **Antibiotic outcomes** |  |  |  |  |  |
| Total DDD received |  |  |  |  |  |
| Median (IQR) | | 3.0 (0.3-6.3) | 6.8 (3.6-10.4) | 4.2 (1.3-8.3) | p<0.001<sup>\$</sup> |
| Total Antibiotic DOT |  |  |  |  |  |
| Median (IQR) | | 2 (0-5) | 5 (4-9) | 5 (1-7) | p<0.001<sup>\$</sup> |
| DDD received/evaluable day |  |  |  |  |  |
| Median (IQR) | | 0.14 (0.02-0.31) | 0.37 (0.19-0.76) | 0.23 (0.08-0.48) | p<0.001<sup>\$</sup> |
| Days of treatment per evaluable day to day 28 |  |  |  |  |  |
| Median (IQR) | | 0.11 (0.00-0.25) | 0.32 (0.18-0.60) | 0.18 (0.04-0.39) | p<0.001<sup>\$</sup> |
| **Infective complications** |  |  |  |  |  |
| HAP/VAP | No | 190 (87) | 127 (85) | 317 | p=0.497<sup>*</sup> |
|  | Yes | 28 (13) | 23 (15) | 51 |  |
| ESBL/AmpC isolation | No | 213 (98) | 148 (99) | 361 | p=0.705<sup>#</sup> |
|  | Yes | 5 (2) | 2 (1) | 7 |  |
| MRSA | No | 218 (100) | 149 (99) | 367 | p=0.408<sup>#</sup> |
|  | Yes | 0 (0) | 1 (1) | 1 |  |
| CDAD | Yes | 0(0) | 0(0) | 0(0) |  |

### Antibiotic Usage

Data on total DDD of antibiotics received in the 28-day follow-up period and DDD per evaluable day are presented in Table 2 and Figure 2. Patients with a negative PCT received significantly fewer DDDs of antibiotics (both total and per evaluable day) than those with positive PCT with a median DDD of 3.0 vs 6.8 (p<0.001). Due to the log-normal distribution of total DDD, a log transformation was performed on the values (after adding the smallest non-zero value of 0.17 to ensure patients with a total DDD of zero were included). Therefore, a log-linear model was computed in order to explore the relationship with PCT positivity after adjusting for demographic confounders (comorbidities, age, sex, ethnicity) to ensure regression assumptions were met. A statistically significant relationship between PCT and total DDD remained after accounting for these confounders; on average a person with PCT>0.25 had almost three times as many DDDs of antibiotics compared to those ≤0.25 (coefficient 2.72, 95%CI: 2.03, 3.62, p<0.001) (Supplementary Table 1).

**Figure 1:**
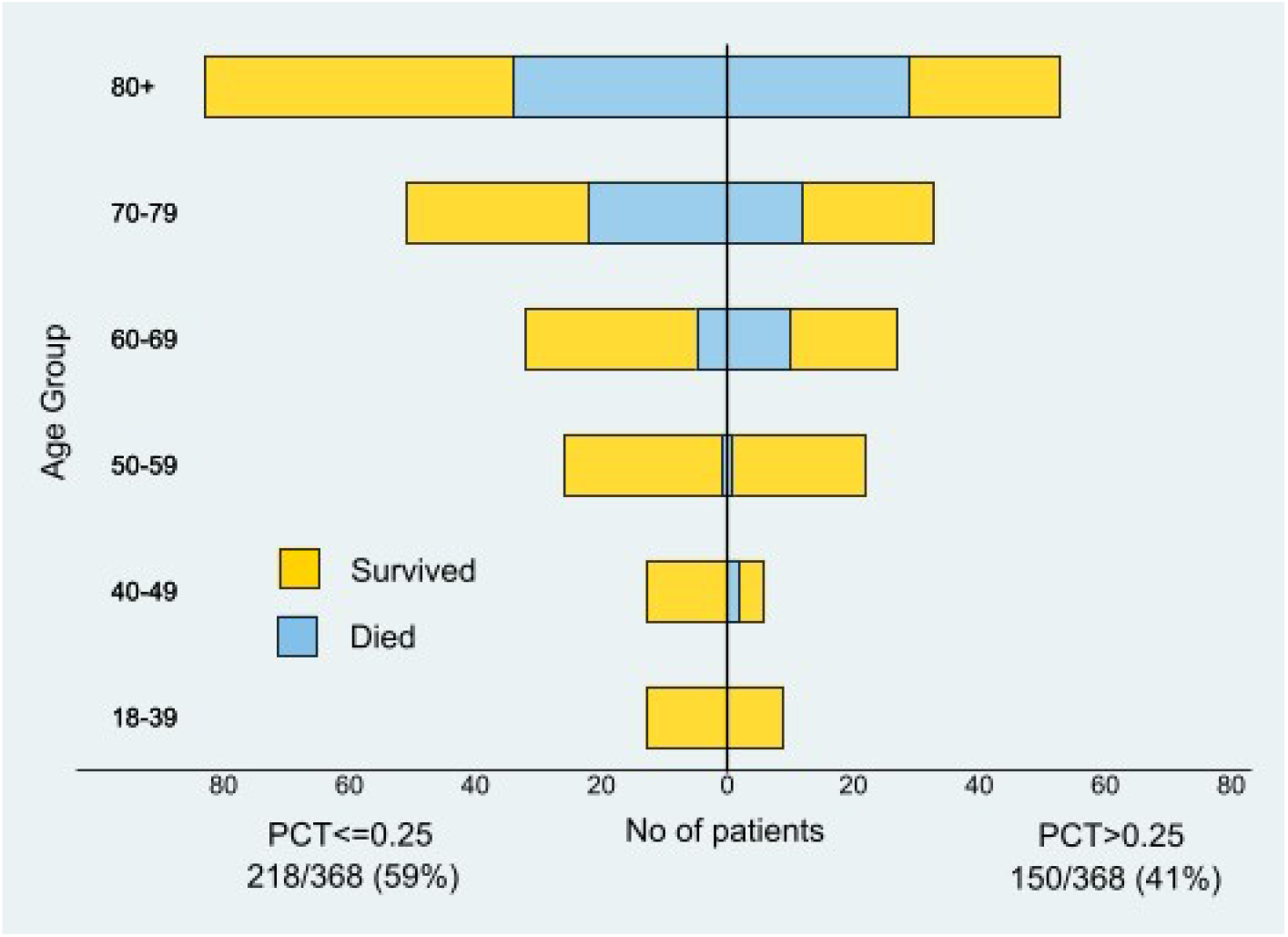
Mortality outcomes for positive and negative PCT groups stratified by age.

**Figure 2.**
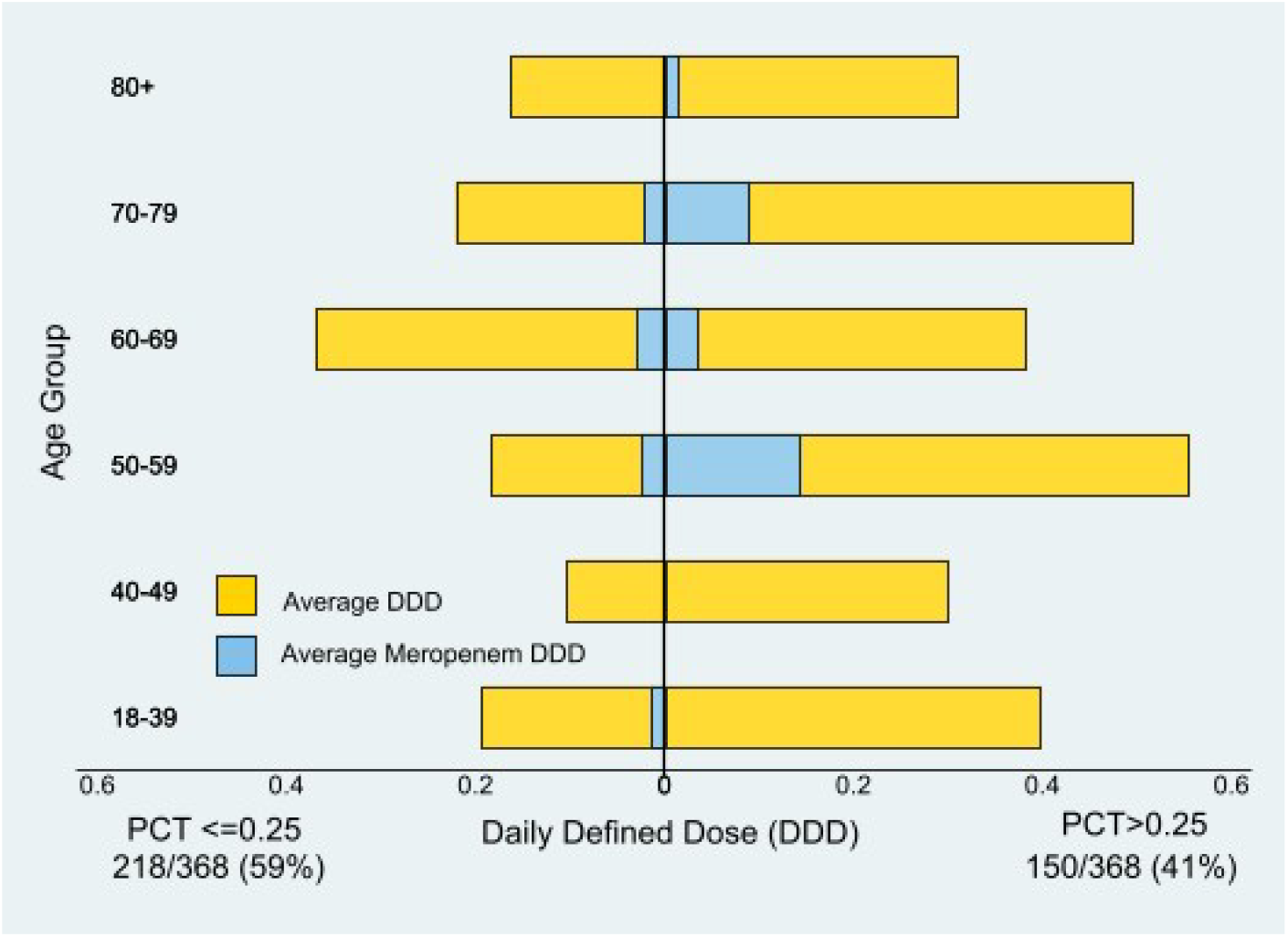
Antibiotics consumption as demonstrated by average DDD & average meropenem DDD between positive and negative PCT groups, stratified by age.

### Patient 28-Day Outcomes

Over the 28-day follow-up period, 116 (32%) of the included patients died, 229 (62%) were discharged and 23 (6%) were still in hospital. Median length of stay was 8.35 days. 47 (13%) were admitted to intensive care, and of these, 32 (68%) were intubated and ventilated. The PCT, age and 28-day mortality distribution of the patients are illustrated graphically in Figure 1. In the PCT negative group, 62 (28%) patients died compared to 54 (36%) of those with a positive PCT (p=0.021), and 19 (9%) were admitted to ITU, compared with 28 (19%) of the positive PCT group (p=0.007); Table 2.

Meropenem was the only carbapenem used in the study population. With specific reference to meropenem consumption, positive PCT was associated with a 3-fold increase in the odds of receiving any meropenem during the course of the admission (OR= 3.16, 95% CI: 1.50, 6.65, p=0.002) after adjusting for demographic confounders (Supplementary Table 2). There was also a statistically significant association between receipt of antibiotics at 48 hours after COVID-19 diagnosis and receipt of meropenem (OR=3.63, 95% CI: 1.53,8.63, p=0.003). (Supplementary Table 3).

Data on the potential adverse consequences of antibiotic usage are presented in Table 2. With the exception of hospital or ventilator associated pneumonia, adverse events were all uncommon and none were significantly different between PCT positive and negative groups.

## DISCUSSION

This observational study supports the hypothesis that implementation of a local guideline advising against the use of antibiotics for patients with confirmed COVID-19 and a PCT level ≤0.25, leads to reduced antibiotic consumption without negative impact on patient 28-day outcome. 28-day mortality figures in this study (28% PCT ≤ 0.25, 36% PCT > 0.25) are similar to data published by the ISARIC consortium, the largest COVID-19 patient registry in the UK, in which 7080 of 19983 (35.4%) patients for whom outcome data was available died [5]. To our knowledge, this is the first study reporting data from the use of PCT as an antimicrobial stewardship tool in patients with COVID-19 and supports its use in this context.

Previous studies have demonstrated that procalcitonin-guided therapy in lower respiratory tract infections substantially reduces antibiotic use without compromising outcome [22-24], including in the critically ill [25]. Although some studies have excluded immunocompromised patients when evaluating PCT, others have shown that a rise in the biomarker correlates with bacterial infection [26, 27]. Drawbacks of PCT-guided therapy include the potential for false negative results in localised infection such as empyema, in atypical infection – and in the context of renal replacement therapy [28, 29]. As it may take 24-48 hours for PCT to reach its peak, false negatives may be seen if samples are taken early in the course of infection [30, 31]. PCT levels also fall by 50% every 24-36 hours [31] and false negatives may occur in patients due to resolution of infection. It is important, therefore, that interpretation of PCT is made in the context of other laboratory and clinical findings [17].

PCT results can be obtained rapidly from a readily obtained sample. This presents a major advantage over respiratory tract culture results, which are challenged by the inability to obtain adequate sample, slower turnaround times and insensitivity. Sputum culture has been shown to reveal a definitive diagnosis in less than 20% of cases of community-acquired pneumonia [32]. Clinical uncertainty, combined with this diagnostic insensitivity, further compromised following initiation of antibiotics, has the potential to provoke unnecessary antibiotic usage and consequent short- and long-term morbidity in patients without bacterial infection.

The adopted PCT threshold of 0.25 was intentionally conservative and it may be that a higher threshold can be adopted safely, which would lead to a further reduction in antibiotic usage in patients with COVID-19.

This higher mortality seen in the PCT >0.25 group supports those of other authors, demonstrating an association between higher PCT values and severe disease or death [33, 34]. It is possible that higher PCT in these patients reflects bacterial superinfection, which increases the production of PCT from non-thyroidal sources through the interplay of interleukin (IL)-1β, tumor necrosis factor (TNF)-α and IL-6, whereas viral infections result in a rise in interferon (INF)-γ which inhibits PCT synthesis [16]. It is also possible that PCT is raised in severe COVID-19 disease independent of bacterial infection, which would open the possibility of further improvements in antimicrobial stewardship.

Many of the advances in medicine that have occurred over the past 75 years have been made possible because of the effectiveness of antibiotics to prevent or treat infective complications. Antimicrobial resistance (AMR) poses a major threat to human health and, if unchecked, it is estimated that 10 million deaths may be attributable to AMR by 2050.

Reducing the unnecessary use of antibiotics is a key component to mitigating this risk. The risk of severe COVID-19 disease increases with age and the elderly are also at greatest risk of the adverse consequences of excessive antibiotic use [35].

Numerous metrics exist for the measurement of antibiotic consumption, each with their own strengths and weaknesses [21]. World Health Organisation Defined Daily Doses have the advantage that they are standardised and, at institution level, can be assessed from procurement records without recourse to individual prescribing data. They do, however, have several drawbacks. They do not factor in exposure equivalent dose adjustments such as those that occur in renal or liver dysfunction, and combination therapy results in elevation of the DDDs received which may correlate poorly with relative effects on host ecology and adverse consequences of therapy. Finally, they are based on usual prescribing patterns and therefore markedly different DDD may apply to different settings or routes of administration of the same drug [36]. This latter weakness was addressed in our study through the use of the parenteral DDD for those drugs where there was a difference. We also assessed days of treatment (DOT), which may be a more relevant metric from a stewardship perspective and again this was found to be significantly lower in the negative PCT group.

We also demonstrated a 3-fold increase in the odds of carbapenem prescription in those with a positive PCT. This is important in the context of the increasing global incidence of carbapenemase-producing Enterobacteriales and is likely to be a direct consequence of increased initial antibiotic usage and a concern that this had induced resistance leading to prescription of broader spectrum agents for any subsequent indication and despite the fact that isolation of ESBL or AmpC positive organisms occurred no more frequently in this group. The impact of early antimicrobial therapy on later prescription of broad spectrum agents is supported by the association between antibiotic use at 48 hours and subsequent meropenem use, which was also statistically significant.

The limitations of our study include the fact that it is from a single centre and retrospective in design. However, patients were recruited systematically from the beginning of the epidemic locally until a date which was defined in advance and which preceded the introduction of routine COVID-19 testing of all admissions regardless of symptoms. It is possible that some patients may have been readmitted to other regional hospitals, but this is felt unlikely as the catchment area of STHNFT is well-defined and there is limited geographical overlap with neighbouring trusts.

## CONCLUSIONS

This study has demonstrated that procalcitonin level can be a useful tool in the assessment of bacterial co-infection of COVID-19 patients, and can safely reduce unnecessary antibiotic usage and its associated adverse consequences.

## Data Availability

The patient data referred to in the manuscript was retrospectively collected by our team and is outlined within the manuscript itself and in the supplementary materials.

## ACKNOWLEDGEMENTS

The authors would like to thank Dr Eirini Koutoumanou of UCL for discussion around the transformation analysis.

